# A New Differential Gene Expression Based Simulated Annealing for Solving Gene Selection Problem: A Case Study on Eosinophilic Esophagitis and Few Other Gastro-Intestinal Diseases

**DOI:** 10.1101/2024.05.03.24306738

**Authors:** Koushiki Sinha, Sanchari Chakraborty, Arohit Bardhan, Riju Saha, Srijan Chakraborty, Surama Biswas

## Abstract

**Background:** Identifying the set of disease-causing genes is crucial for understanding pathogenesis and developing therapies. This is particularly important to understand the pathophysiology of Eosinophilic Esophagitis (EoE) and other gastrointestinal diseases. Comparing and contrasting gene selection methods across these diseases can enhance our knowledge to identify potential therapeutic targets.

**Methods:** This study introduces two approaches for gene selection in gastrointestinal diseases: the Ranked Variance (RV) method and Differential Gene Expression Based Simulated Annealing (DGESA). RV acts as an initial screener by prioritizing genes based on variance. DGESA refines gene selection further by employing simulated annealing with differential expression data. We compared the outcomes of both methods through a case study on EoE and other gastrointestinal diseases.

**Results:** Result finds greater number of genes with negative fold changes compared to those with positive fold change in differential EoE dataset. RV Ranks top 40 genes with high variance of EoE which overlaps with the disease-causing gene set of EoE from DGESA. 40 gene pathways for each of EoE, Crohn’s Disease (CD), and Ulcerative Colitis (UC) were identified as execution outcome of our method DGESA. Among these, 10 genes for EoE, 8 for CD, and 7 for UC were confirmed in the literature for their connection with respective diseases. For EoE, 10 such confirmed genes include KRT79, CRISP2, IL36G, SPRR2B, SPRR2D and SPRR2E. For CD, the literature confirmed set encompasses NPDC1, SLC2A4RG, LGALS8, CDKN1A, XAF1, and CYBA. The validated genes in UC final gene set includes TRAF3, BAG6, CCDC80, CDC42SE2, and HSPA9.

**Conclusion:** The RV method, serving as an initial screener, and the more refined DGESA both effectively elucidate molecular signatures in gastrointestinal diseases. Identifying and validating genes like SPRR2B, SPRR2D, SPRR2E and STAT6 for EoE showcase efficacy of DGESA. Other genes in the same pathway are interesting targets for future laboratory validation.

## Introduction

Computational Genomics is one of the most emerging areas of biological research. It is empowered by the fusion of powerful computational algorithms and vast genomic datasets. Its pivotal role in answering complex biological questions is evident in its application across diverse domains, from tracing the origins of diseases like SARS to provide insights on cancer immunotherapy [1]. By utilizing statistical methods and computational tools, bioinformaticians decode the language of genomes, offer understanding of the evolutionary mechanisms, genomic variations and gene interactions. Recent strides in high-throughput sequencing technologies have amplified the volume and complexity of genomic data, necessitating sophisticated computational frameworks for data processing, analysis, and visualization. These frameworks enable researchers to mine large datasets effectively, discovering novel mechanistic insights into fundamental biological processes [2]. Moreover, as evidenced by the publicly available genomic databases and freely downloadable program suits like Genomic Multi-tool (GEM), the democratization of computational tools fosters collaboration and innovation in the scientific community. Computational Genomics not only shades lights on the genetic factors of diseases but also escalates advancements in personalized medicine, drug discovery, and agricultural resilience to climate change [3]. As universities introduce interdisciplinary courses to train the next generation of bioinformaticians, the field continues to evolve, embracing emerging technologies like Machine Learning, Artificial Intelligence and other modern computational methodologies. Through continuous refinement of computational methodologies and the integration of diverse datasets, Computational Genomics promises to revolutionize our understanding of life’s fundamental processes, driving forward the frontiers of both biological discovery and technological innovation [4].

Understanding biological processes requires an understanding of gene expression data, which provide insight into how genes function within cells. There are many public repositories available. A notable instance of such a database is the Gene Expression Omnibus (GEO) from National Center for Biotechnological Information (NCBI). GEO provides high-throughput gene expression and functional genomics datasets in a consolidated manner, making important information accessible to researchers across the globe. Many genomic data categories, such as, chromatin structure, genome methylation, genome-protein interactions and gene expression studies are gathered in this database [5]. GEO guarantees the availability of raw data and metadata by following community-driven reporting standards, which promotes reliable research outputs. Scholars employ GEO’s extensive collection to investigate a range of biological inquiries, capitalizing on its intuitive interface and web-based instruments to facilitate streamlined data retrieval, illustration, and examination. Scientific advancement and creativity are fueled by gene expression data from repositories like GEO [6], which is used to drive discoveries in domains including developmental biology, cancer research, and personalized medicine.

Gastrointestinal diseases encompass a diverse array of conditions affecting the digestive tract, ranging from inflammatory bowel diseases like Crohn’s disease and ulcerative colitis to eosinophilic esophagitis (EoE) [7]. EoE, characterized by chronic immune-mediated inflammation of the esophagus, has emerged as a prominent source of upper gastrointestinal morbidity in recent years. With an estimated prevalence of 34.4/100,000 in Europe and North America, EoE presents symptoms such as esophageal strictures, dysphagia, and food impaction, affecting both children and adults [8]. Unlike other conditions associated with esophageal eosinophilia, EoE diagnosis requires symptoms of esophageal dysfunction alongside esophageal biopsies demonstrating at least 15 eosinophils per high-power field. Genetic and environmental factors, including early exposure to antibiotics, are implicated in its etiology. Current treatment modalities for EoE include proton pump inhibitors, dietary therapy, topical steroid formulations, and endoscopic dilatation, tailored to individual patient needs and disease severity. As our understanding of EoE evolves, further research into its pathogenesis, natural history, and optimal management strategies remains essential for improving patient outcomes and quality of life [9-11].

Genetic factors play a significant role in the pathogenesis of gastrointestinal diseases, with Eosinophilic Esophagitis (EoE) standing out as a prime example. EoE, a chronic allergic condition characterized by eosinophilic infiltration of the esophageal mucosa, is influenced by both hereditary and environmental factors [12]. Studies have highlighted the substantial familial component of EoE, with evidence suggesting a higher likelihood of the condition in family members of affected individuals [15]. Environmental risk factors also contribute to modulating genetic risk in EoE, particularly through early-life events [13]. While rare genetic variations may account for a small subset of EoE cases, the majority of genetic risk is mediated by common genetic variations [15]. Genome-wide association studies (GWAS) have identified specific risk loci associated with EoE susceptibility, such as variants in genes like TSLP and CAPN14, shedding light on the molecular mechanisms underlying the disease [14]. Interestingly, many of these risk loci are located in non-coding regions of the genome, suggesting a role for gene regulation in EoE pathogenesis [15]. Understanding the genetic architecture of EoE not only enhances our comprehension of its molecular basis but also holds promise for the development of targeted therapeutic interventions and personalized treatment strategies based on individual genetic profiles [14]. Further research into the intricate interplay between genetic predisposition, environmental triggers, and immune dysregulation is essential for advancing our understanding of EoE and improving patient care [12].

The prevalence of eosinophilic esophagitis (EoE) in the Indian population appears to be gradually gaining recognition, although with limited data available. A study conducted in a hospital in the northern region of India reported a prevalence of 3.2% among patients with symptoms suggestive of gastroesophageal reflux disease (GERD) [16]. Similarly, another study noted an increase in diagnosed cases of EoE in their center over recent years, with 17 out of 73 patients being diagnosed with EoE based on clinical, endoscopic, and histopathologic features [17]. These findings suggest that EoE exists in India and is gaining importance of clinical suspicion and diagnostic evaluation for its identification. However, the prevalence of EoE in the Indian population remains to be fully explained, highlighting the necessity for large-scale, multi-centric population-based studies to provide a more comprehensive understanding of the disease burden in the country [17].

Gene selection, a critical task in gene expression studies, aims to identify subsets of genes that are relevant for distinguishing between disease and normal conditions. Traditional methods for gene selection often face challenges such as high within-class variation and the selection of an optimal subset of genes that can efficiently differentiate between classes. Recent advancements in artificial intelligence (AI) and machine learning (ML) have introduced novel approaches to address these challenges. For instance, in a study [18], a novel criterion was proposed for assessing the significance of individual genes based on their mean and standard deviation of distances from each sample to the class centroid. This method not only effectively tackles within-class variation but also offers a smaller number of genes without compromising discriminating power, thus supporting further biological and clinical research. Similarly, the utilization of ML techniques, such as random forest, has been demonstrated to be effective in gene selection for microarray data classification [19]. Random forest excels in handling large numbers of variables and noisy data, providing accurate predictions while simultaneously offering small sets of genes for classification. Additionally, machine learning-based approaches have been employed to enhance the stability of gene selection techniques under sample fluctuations. For example, a study by [20] introduced a framework of sample weighting to increase the stability of representative feature selection algorithms, leading to the identification of more stable gene signatures. Furthermore, the application of ML techniques extends beyond gene selection to various aspects of cancer research, including cancer subtype classification, prognosis prediction, and identification of biomarkers. Studies such as [21, 22, 23, 24] highlight the utility of ML algorithms like XGboost and support vector machine (SVM) in classifying cancer subtypes and identifying potential biomarkers for hepatocellular carcinoma (HCC) and Crohn’s disease (CD), respectively. These approaches leverage large-scale genomic data to facilitate personalized treatment strategies and improve patient outcomes. In summary, the application of AI and ML techniques holds great promise in addressing the gene selection problem by providing efficient, accurate, and stable methods for identifying disease-relevant genes and advancing our understanding of complex diseases [25, 26, 27].

Gene expression variance plays a crucial role in computational genomics, influencing our understanding of complex genetic diseases and population genetics. Studies such as [28] have highlighted the significance of genetic variants in Alzheimer’s disease (AD) risk, with known single nucleotide polymorphisms (SNPs) explaining only a portion of the phenotypic variance. This represents the importance of exploring additional sources of genetic variation, such as rare or unknown SNPs, to take into account the full genetic landscape of AD. Similarly, research on the ALDH2 gene variant, as discussed in [29], sheds light on how specific genetic variations can influence susceptibility to various diseases and physiological traits. Understanding the mechanisms underlying these associations is essential for precision medicine and disease prevention strategies. Furthermore, investigations into gene expression variance, as described in [30], provide insights into the regulatory mechanisms governing gene expression across different tissues and conditions. By observing patterns of transcriptional variance and its relationship with gene function, computational genomics can identify key regulatory elements and pathways underlying complex traits and diseases. Overall, the study of gene expression variance in computational genomics enhances our understanding of genetic diversity, disease susceptibility, and molecular mechanisms, paving the way for more effective diagnostic and therapeutic interventions.

Simulated Annealing (SA) draws inspiration from metallurgy’s annealing process, where material is gradually cooled to a stable state. Introduced by Kirkpatrick, Gelatt, and Vecchi [31], SA optimizes by navigating a solution space, occasionally accepting moves that increase the cost function value (in minimization). As it progresses, the algorithm gradually reduces the likelihood of accepting worse solutions, akin to cooling. SA’s extensions include adaptations to combinatorial optimization and nonconvex problems [32-33]. In genomic feature selection, SA aids in identifying relevant genes from high-dimensional datasets, such as microarray gene expression data. Recent advancements like curious simulated annealing [34] and Simulated Annealing aided Genetic Algorithm (SAGA) [35] address SA’s convergence limitations. They strike an optimal balance between exploring the search space and exploiting potential solutions.

The gene selection problem, which focuses on identifying a set of genes that collectively cause a disease, is crucial for understanding complex medical conditions. Though many complex formulations are already available, a very simple, efficient and biologically plausible fact that optimization process needs guidance from the differentiability of diseased to normal genomic profiles was overlooked. This study introduces a new Simulated Annealing based algorithm called Differential Gene Expression Based Simulated Annealing (DGESA) where a specially designed objective function has been introduced which aims to maximize the collective differentiability of the obtained genes in their diseased and normal genomic profiles. For an initial guess of differentiability in the gene expression data, an approach, termed here as Ranked Variance (RV) has been introduced that prioritize genes based on their variance. Through a case study on Eosinophilic Esophagitis (EoE) and other gastrointestinal diseases, we compare the outcomes of both methods. Notably, we find 10 common genes between RV and DGESA in EoE, indicating their complementary nature. Validation analyses show that 10 of the 40 final genes identified by DGESA for EoE are supported by existing literature, confirming their biological relevance. Similarly, for Ulcerative Colitis (UC) and Crohn’s Disease (CD), 8 and 7 of the 40 genes, respectively, are validated by literature. Ten confirmed genes for EoE are as follows: KRT79, CRISP2, IL36G, SPRR2B, SPRR2D, and SPRR2E. The collection of CD that has been confirmed by literature includes NPDC1, SLC2A4RG, LGALS8, CDKN1A, XAF1, and CYBA. The final gene set from UC contains the validated genes TRAF3, BAG6, CCDC80, CDC42SE2, and HSPA9. These results underscore the efficacy of our framework and specifically DGESA in identifying significant molecular signatures associated with gastrointestinal diseases.

## Methodology

The method section of this study details two distinct approaches employed for gene selection and analysis: the RV method and DGESA. The RV method prioritizes genes based on their variance, providing an initial perspective on potential biomarkers. In contrast, DGESA utilizes simulated annealing to identify sets of genes exhibiting significant differences in expression between diseased and normal states, facilitating the discovery of disease-associated genetic signatures. Each method offers unique insights into gene selection and contributes to our understanding of molecular mechanisms underlying disease pathogenesis (see Figure 1).

**Figure 1:**
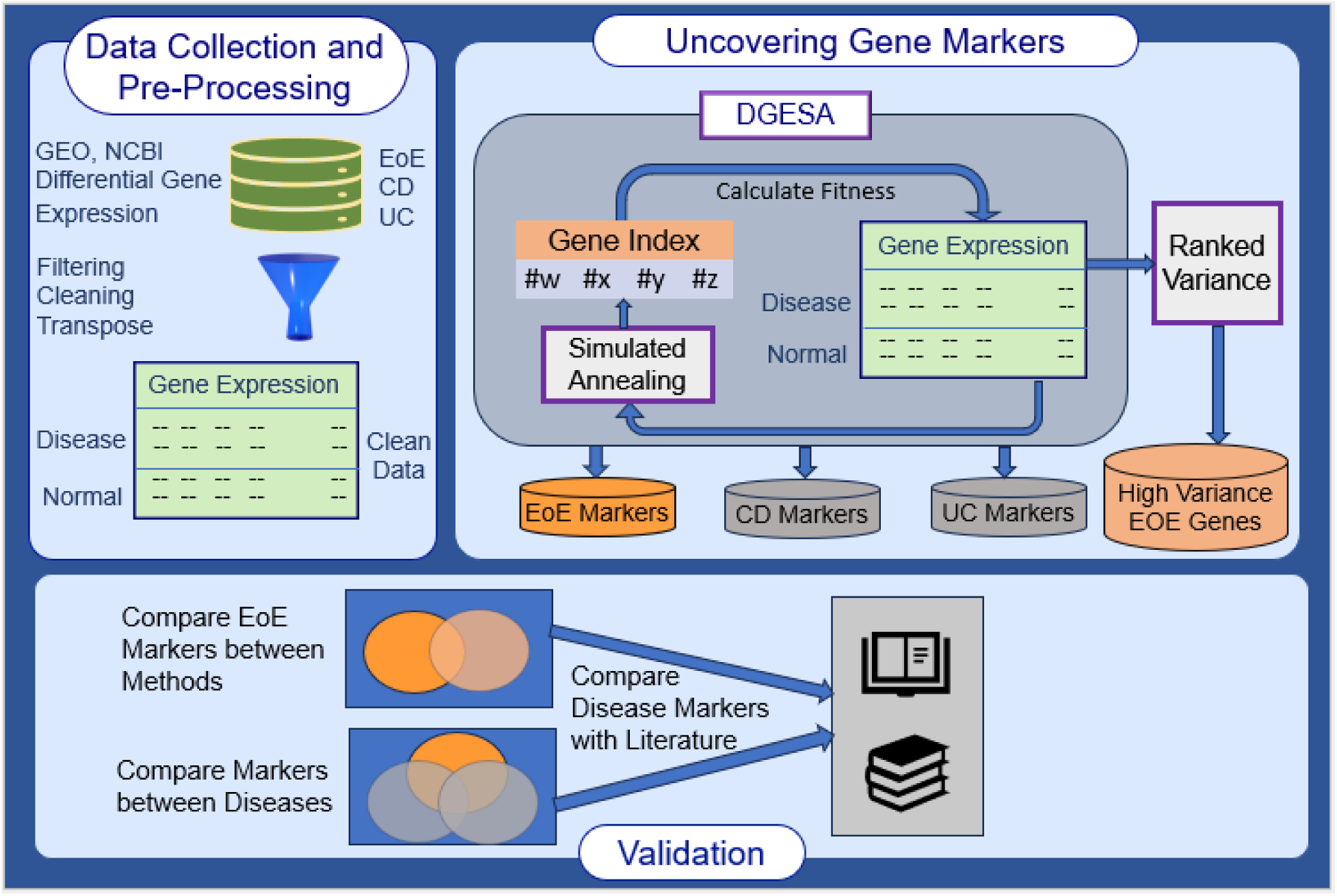
Overview of the Study Workflow. This figure illustrates the comprehensive workflow of the study, beginning with data acquisition and preprocessing steps that include cleaning, normalization, and transposition to produce the gene expression matrix (samples as rows, genes as columns). The gene marker identification methodologies encompassing RV and DGESA, are applied to datasets of EOE, CD, and UC to identify disease-specific gene markers. The final step involves the validation process, which compares the effectiveness of the two methods across the three diseases and validates the identified markers against existing literature.

**Figure 2:**
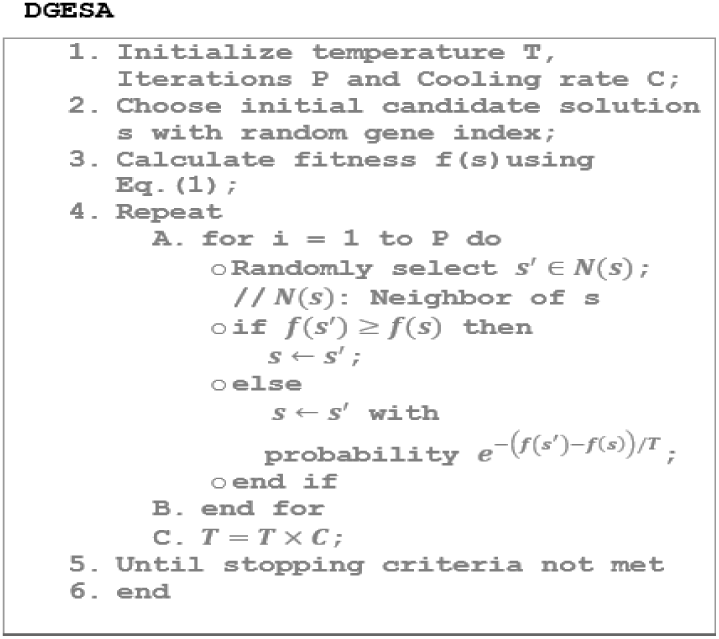
DGESA Algorithm. This figure depicts the details of the DGESA algorithm. The process involves simulated annealing with a specially designed objective function to effectively select gene markers from differential gene expression data.

Prior to analysis, several preprocessing steps were implemented to ensure data quality and compatibility. Firstly, rows lacking valid gene names were removed to maintain consistency across datasets. Subsequently, a normalization procedure was applied to each dataset, wherein gene expression values (𝑒_𝑖,𝑗_) were mapped to the range [0, 1]. This normalization step helped maintain potential biases arising from variations in gene expression magnitude across samples. Finally, transposition of the datasets was performed to prepare the data matrix (denoted as X) for subsequent processing, facilitating the application of required analytical techniques. These preprocessing steps collectively ensured that the gene expression data were standardized and conducive to meaningful analysis and interpretation.

The Ranked Variance (RV) method employed in this study focused on using gene expression variability as a means of discerning potential biomarkers associated with disease states. By computationally analyzing the variance of gene expression across samples, the RV method identified genes exhibiting significant variations in expression levels. This approach facilitated the separation of disease-associated genes from those with relatively stable expression patterns, thereby providing valuable insights into the molecular mechanisms underlying disease pathogenesis. Moreover, the identification of genes with pronounced expression variations enabled subsequent association studies, wherein these genes could be further investigated for their roles in disease development, progression, and potential therapeutic targeting. Overall, the RV method served as a powerful tool for examining the genetic signatures associated with various diseases, contributing to our understanding of their underlying biological processes and aiding in the discovery of novel biomarkers.

DGESA is a methodology devised to address gene selection challenges in the context of biological diseases. At its core, DGESA operates on a transposed gene expression matrix (denoted as X), where each row represents an observation (diseased or normal person) and each column corresponds to a gene. The method begins by defining a candidate solution, represented as a set of gene indices, which is iteratively refined through a simulated annealing process. During each iteration, a perturbation is applied to the current solution by randomly altering a gene index from the gene expression matrix X that is not already present in the solution set s. The fitness of each candidate solution is evaluated using a devised fitness function, represented by Equation (1):

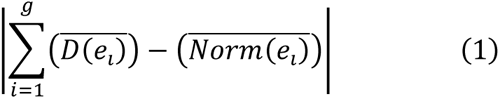

Here, g represents the number of genes in the candidate solution s. For each gene index *i* in *s*, the expression profile (𝑒_𝑖_) is considered. The mean expression profile of the *i*-th gene in the candidate solution s for diseased samples is denoted by 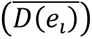, while the mean expression profile for normal samples is denoted by 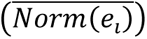. The fitness function computes the absolute difference between the mean expression profiles of diseased and normal samples across all genes in the candidate solution. This difference serves as a measure of the discriminative power of the selected genes in distinguishing between diseased and normal states.

In DGESA, the Temperature (𝑇) is first initialized into a very high value. Then few simulated annealing steps have been performed until the stopping criteria are not met. In each such step, 𝐿 neighborhood search iterations, followed by a reduction of 𝑇 by a fraction of Cooling Factor 𝐶 have been performed. In each neighborhood search state, we select a neighbor 𝑠^′^ from the neighborhood of s and replace 𝑠 by 𝑠^′^ if 𝑠^′^is better than 𝑠 in terms of fitness value otherwise 𝑠 may be replaced by 𝑠^′^ depending on a small probability. Through this iterative optimization process, DGESA aims to identify a set of genes that collectively exhibit significant differences in expression patterns between diseased and normal samples. The output of DGESA, denoted as 𝑠^∗^, represents the final selection of genes maximizing the discrimination between diseased and normal conditions in gene expressions, thereby facilitates the identification of potential biomarkers and provides insights into disease mechanisms.

The methods applied in this study, including RV method and DGESA, have provided valuable insights into gene selection and analysis within the context of gastrointestinal diseases. The RV method effectively identified genes with significant expression variations, aiding in disease gene separation and association studies. On the other hand, DGESA upgrades simulated annealing to pinpoint genes exhibiting differential expression patterns between diseased and normal samples, thereby contributing to the discovery of disease-associated genetic signatures. By employing these complementary methodologies, the understanding of molecular mechanisms underlying disease pathogenesis have been advanced and a robust framework for biomarker discovery and disease classification is presented.

## Results

The data collection for this study involved retrieving two gene expression datasets from GEO, NCBI. The first dataset, GSE228083, comprised samples from patients with EoE compared to normal samples, facilitating the investigation of gene expression patterns specific to this condition. The second dataset, GSE24287, encompassed gene expression profiles from patients with UC, CD, and normal samples. From GSE24287, two distinct datasets were prepared by segregating samples into UC vs. Normal and CD vs. Normal categories.

Hyper-parameter tuning of DGESA is a critical aspect of optimizing its performance in gene selection tasks. This iterative process involves systematically adjusting parameters that control the learning process, known as hyper-parameters, through experimentation with different configurations. In the case of DGESA, key hyper-parameters include the number of genes (𝑔) in the candidate solution, the maximum number of iterations, the initial temperature (𝑇), and the cooling rate (𝐶). By systematically adjusting these hyper-parameters, the DGESA model can be fine-tuned to enhance its efficiency in identifying disease-associated genes. In this study, after thorough experimentation and analysis of resulting performance of the algorithm to convergence, the final hyper-parameter configurations were determined as follows: 𝑔 = 40 genes in the candidate solution, 200,000 iterations, T = 10^6^, and a cooling rate of 0.9. These optimized hyper-parameters ensure the effectiveness of DGESA in identifying relevant genetic signatures associated with gastrointestinal diseases, thereby advancing our understanding of disease mechanisms and aiding in biomarker discovery.

To gain insight into the differential expression patterns of genes in the EoE dataset, a volcano plot was generated, depicting the relationship between the log2 fold change and the log10 p-values of various genes. In this plot, the x-axis represents the log2 fold change, which quantifies the magnitude of gene expression differences between EoE samples and normal samples. Meanwhile, the y-axis displays the -log10 p-values, which serve as a measure of the statistical significance of these expression differences. The volcano plot (see Figure 3) revealed that the majority of genes exhibited negative fold changes, indicating under-expression in EoE compared to normal samples. This observation suggests a potential downregulation of gene expression associated with EoE pathology. However, it’s essential to interpret these findings in conjunction with additional analyses to elucidate the specific genes and biological pathways underlying the disease’s pathogenesis and progression.

**Figure 3.**
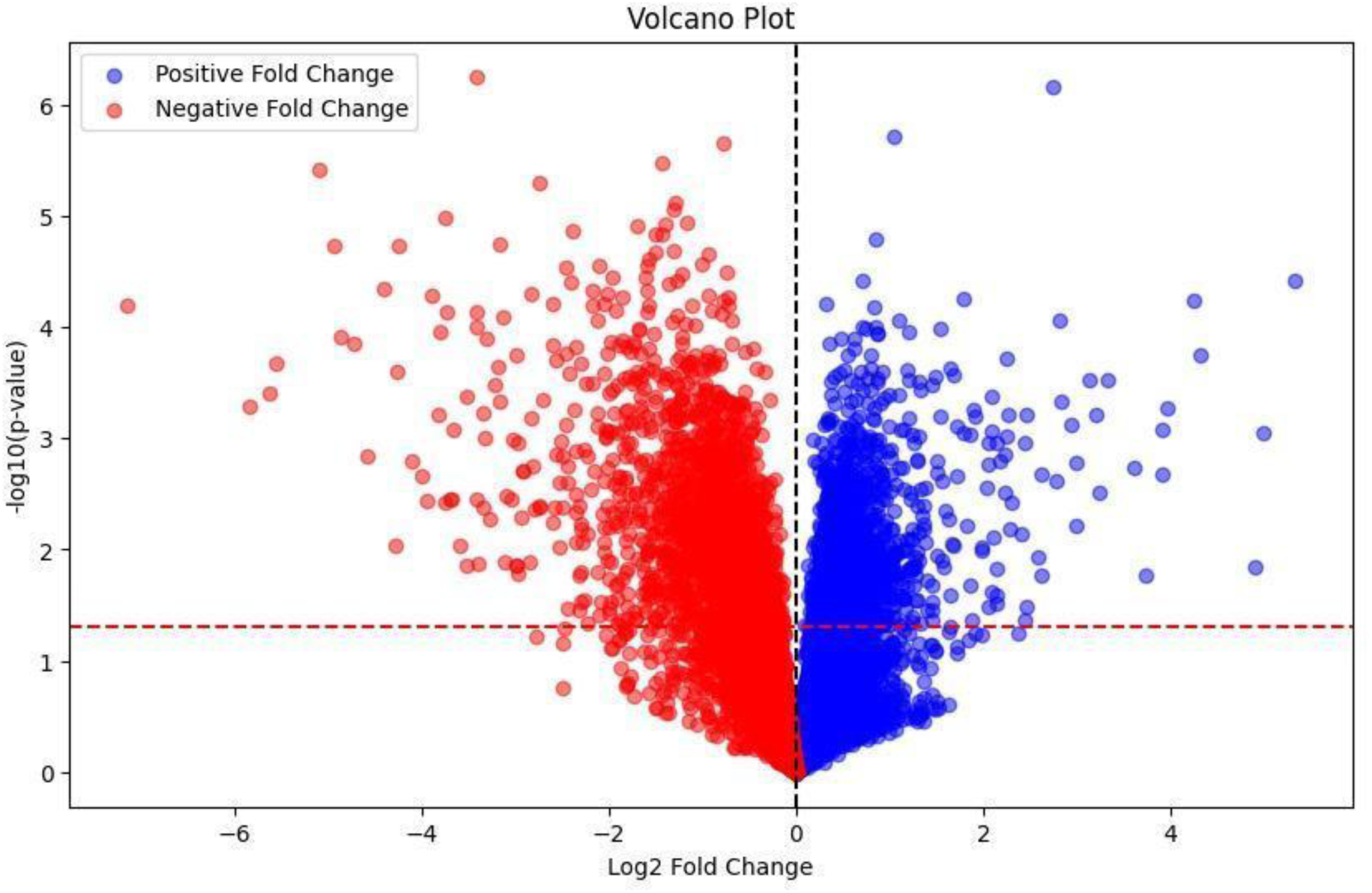
Volcano Plot of Gene Expression Data for EOE: This figure presents a volcano plot illustrating the gene expression data for EOE. The x-axis represents the log2 fold change, while the y-axis denotes the -log10 P-value. The plot highlights a greater number of genes with negative fold changes compared to those with positive fold changes, indicating differential gene expression patterns in EOE.

The application of the RV method to the EoE vs. normal dataset yielded insightful results regarding the variability of gene expression across samples. By plotting the curve (see Figure 4) where the x-axis represents the gene index and the y-axis denotes the corresponding variance, it was observed that approximately 40 genes exhibited decreasing variance. This observation suggests a notable reduction in the variability of expression levels for these genes in EoE samples compared to normal samples. Such a trend of decreasing variance may indicate a degree of regulatory homogeneity or consistent downregulation of gene expression within this subset of genes in the context of EoE pathology. These findings highlight the potential significance of these genes in contributing to the molecular mechanisms underlying EoE development and progression. The variance graph of CD and UC are available on Supplement 1 and 2 respectively.

**Figure 4.**
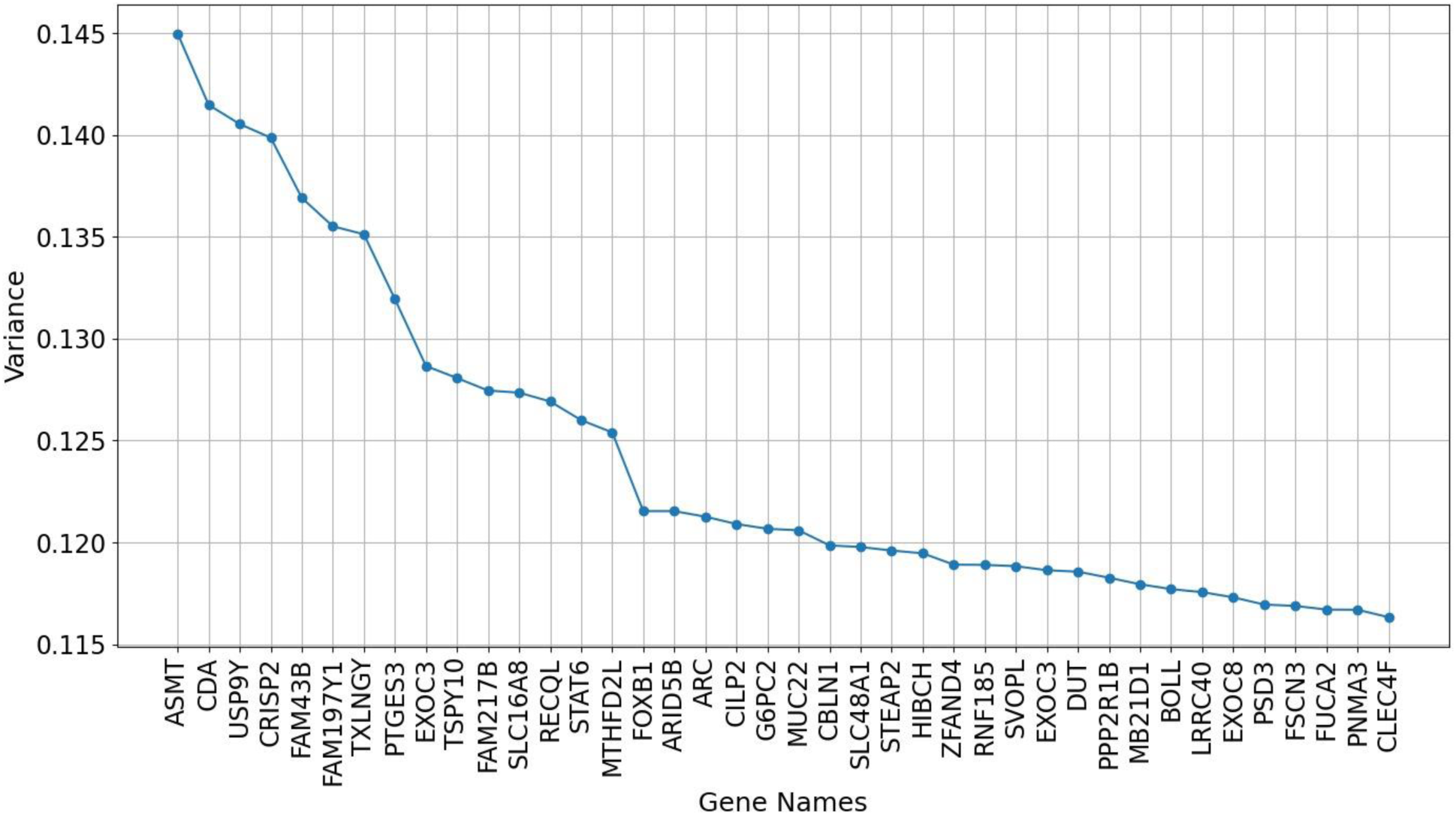
Variance of Top 40 Genes in EOE Gene Expression Data: This figure shows a line plot of the variance of the top 40 genes in the EOE gene expression dataset. The x-axis represents the gene names with the highest variance, and the y-axis indicates the variance values. The plot reveals that the initial few genes exhibit very high variance, which gradually flattens down for the subsequent genes.

The application of the DGESA method to the EoE vs. normal dataset yielded a convergence curve that provides valuable insights into the optimization process. In this curve, the x-axis represents the iterations, reflecting the number of iterations or steps taken during the simulated annealing optimization procedure. Meanwhile, the y-axis denotes the corresponding fitness values, which quantify the effectiveness of the candidate solutions at each iteration. The observed convergence of the curve indicates that as the optimization progresses through iterations, the fitness values gradually stabilize or improve, eventually reaching an optimal or near-optimal solution. This convergence phenomenon signifies the effectiveness of DGESA in iteratively refining the selection of genes to maximize their discriminative power between EoE and normal samples. The convergence curve represents the robustness and efficiency of DGESA in (see Figure 5) identifying disease-associated genetic signatures and highlights its potential as a valuable tool for biomarker discovery. Showcasing the consistency, the optimization curves obtained from CD vs. normal and UC vs. normal datasets show convergence (see Supplement 3 and 4 respectively).

**Figure 5.**
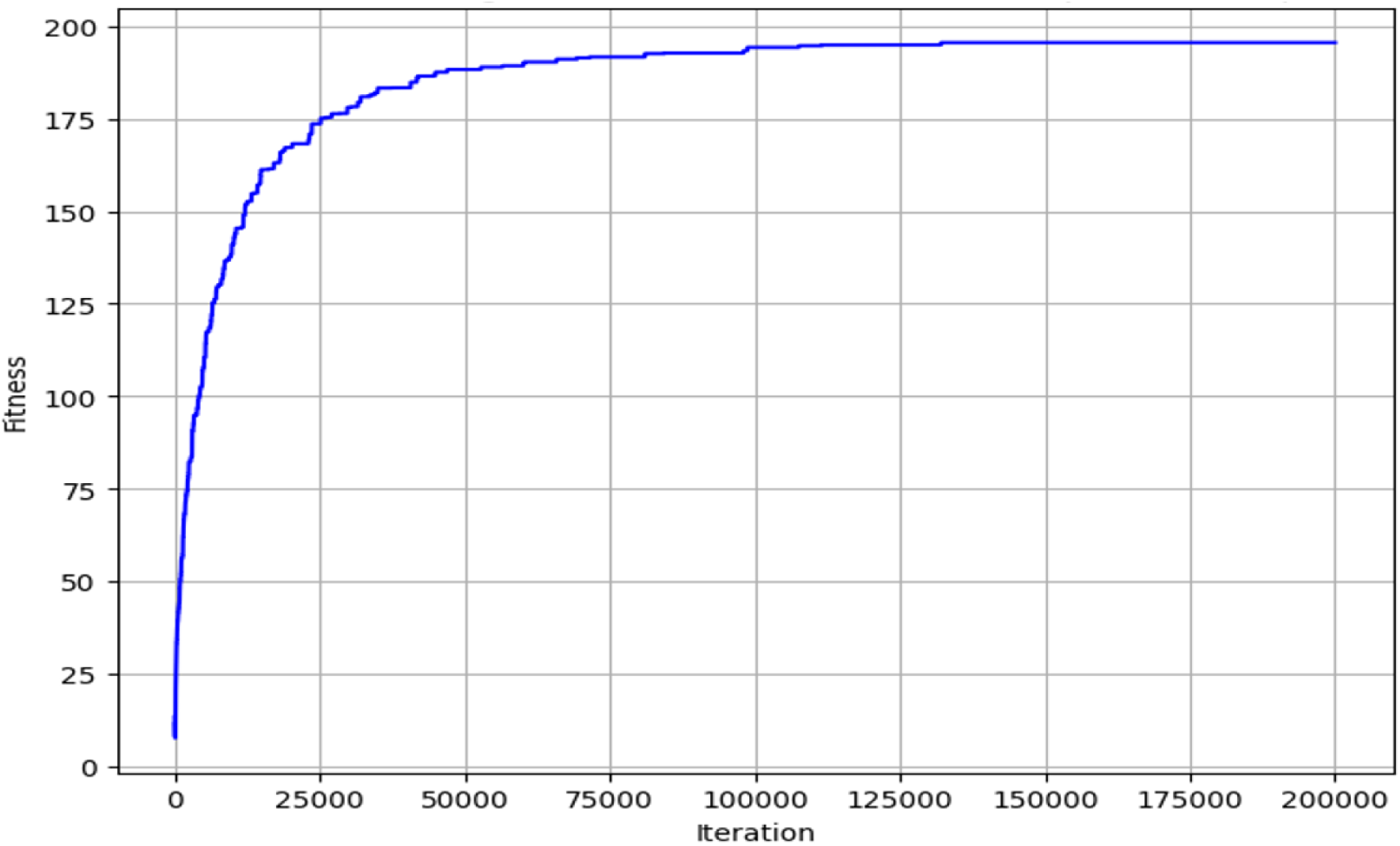
Optimization Process for Identifying 40 Discriminative Genes in EOE: This figure illustrates the optimization process aimed at selecting a set of 40 genes that collectively maximize the discrimination between EOE and control samples in the gene expression dataset. The curve shows the convergence of the optimization process, stabilizing near 130,000 epochs.

The Venn diagram analysis comparing the gene sets identified by the DGESA and RV methods in the context of EoE versus normal samples revealed important findings (see Figure 6). Specifically, the diagram indicated that 10 genes were shared between DGESA and RV (see Table 1 for the common gene names), suggesting a degree of consistency or agreement between the two methods in identifying potential biomarkers associated with EoE. However, notably, no overlap was observed between the gene sets identified for EoE and the combined UC and CD versus normal dataset. This absence of connection between the gene sets for EoE and UC-CD may reflect distinct molecular mechanisms underlying these two gastrointestinal diseases. The lack of shared genes highlights the specificity of gene expression profiles associated with each disease entity and emphasizes the importance of tailored approaches for biomarker discovery and therapeutic targeting. Further investigation into the unique genetic signatures of EoE and UC-CD could offer deeper insights into their pathogenesis and facilitate the development of more precise diagnostic and therapeutic strategies.

**Figure 6.**
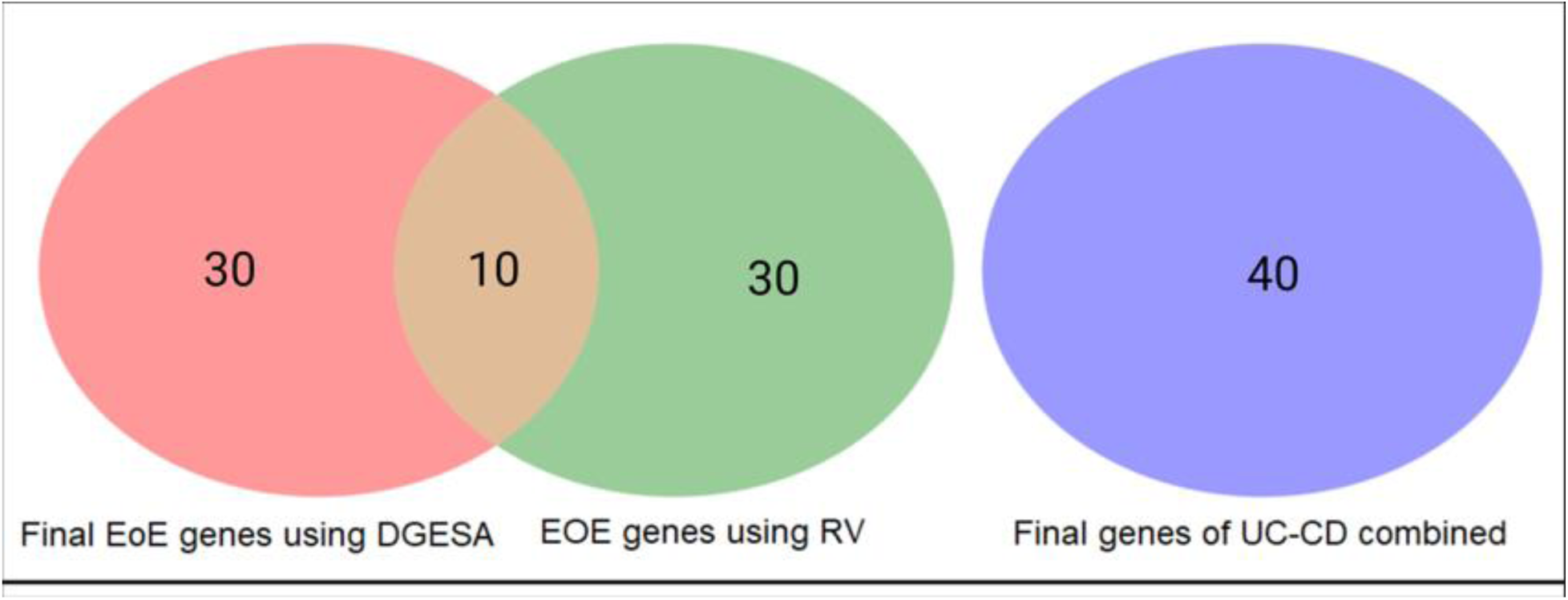
Venn diagram of Common Genes Identified by RV and DGESA Methods in EoE and UC-CD Combined Datasets. This figure shows a Venn diagram comparing the common genes identified by Ranked Variance (RV) and Differential Gene Expression Based Simulated Annealing (DGESA) methods in the EoE gene expression dataset (left side) and the common genes identified using DGESA in the UC-CD combined dataset. The diagram reveals 10 common genes between the RV and DGESA methods for EoE, but no overlapping genes between the EoE dataset and the UC-CD combined dataset.

**Table 1:**
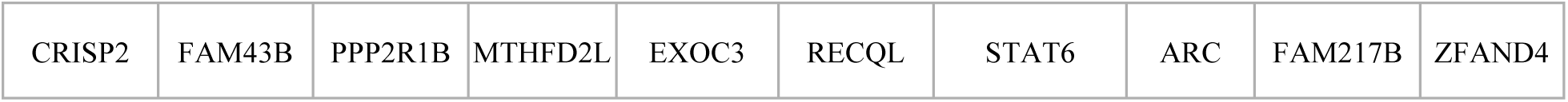
Common genes of EoE obtained using RV and DGESA.

|  |  |  |  |  |  |  |  |  |  |
| --- | --- | --- | --- | --- | --- | --- | --- | --- | --- |
| CRISP2 | FAM43B | PPP2R1B | MTHFD2L | EXOC3 | RECQL | STAT6 | ARC | FAM217B | ZFAND4 |

The final gene set identified by the DGESA algorithm for EoE presents a compelling alignment with previous literature, demonstrating its potential significance in the context of EoE pathology. Among the 40 genes listed (see Table 2), 10 genes, including KRT79 [36], SPRR2E [37], CRISP2 [38], IL36G [39], SPRR2B [37, 40], SPRR2D [37, 40] and RORC [41] have been previously implicated in EoE and are highlighted in blue in Table 2 to denote their strong confirmation of association with the disease. These genes represent key players in various molecular pathways relevant to EoE, such as immune regulation, epithelial barrier function, and tissue remodeling. Interestingly, CCND1 [42] linked with allergy in cow milk, appeared in our result. It might have an indirect link with EoE because cow milk allergy has been considered as one of the factors causing EoE. Additionally, the presence of other genes in the final gene set, although not explicitly highlighted, suggests potential connections to EoE based on their co-appearance with established EoE-associated genes. This comprehensive gene set derived from DGESA not only validates known associations but also offers new insights into the molecular mechanisms underlying EoE pathogenesis, paving the way for further research into diagnostic and therapeutic interventions for this complex disease. The unique genes of CD and UC by DGESA are available on Table 3 and 4 respectively.

**Table 2:** Final unique genes of EoE using DGESA: Here the genes highlighted with blue have confirmed their connection with state-of-the-art literature as related with CD. For example, KRT79 [36], SPRR2E [37], CRISP2 [38], IL36G [39], SPRR2B [37, 40], SPRR2D [37, 40], RORC [41] and CCND1 [42] are confirmed connection with EoE.

|  |  |  |  |  |  |  |  |  |  |
| --- | --- | --- | --- | --- | --- | --- | --- | --- | --- |
| HSPA12A | DPCR1 | FAM25G | FAM43B | IVD | PPP2R1B | MTHFD2L | SPRR2E | GGT6 | KRT79 |
| RORC | CRISP2 | ZNF562 | OAZ3 | C18orf54 | EXOC3 | IL36G | TPPP2 | ANXA8 | CSN2 |
| RECQL | RPAP3 | SPINK13 | TAF4B | LYPD6 | COX6B1 | CPB1 | SPRR2B | SPRR2D | DMKN |
| FAM217B | HIP1 | ARC | ZFAND4 | CCND1 | RMI1 | LOC388780 | CSNK1A1L | ADGRB1 | STAT6 |

**Table 3:** Final unique genes of CD using DGESA: Here the genes highlighted with blue have confirmed their existence in state-of-the-art literature as related with CD. For example, NPDC1 [43], SLC2A4RG [44], LGALS8 [45], CDKN1A [46], XAF1 [47], *CYBA* [48] etc. are confirmed connection with Inflammatory Bowel Syndrome (IBD) like CD.

|  |  |  |  |  |  |  |  |  |  |
| --- | --- | --- | --- | --- | --- | --- | --- | --- | --- |
| EPHX3 | NRG1 | PC | NPDC1 | VTI1A | ZNF646 | TCEA1 | SLC2A4RG | LGALS8 | VDAC3 |
| G2E3 | ATXN1 | KTN1 | PLEK2 | CDKN1A | ACOT8 | GLB1 | XAF1 | CYBA | WDR85 |
| NF1 | ANXA2P1 | PSAT1 | ABHD11 | HPN | PEX19 | MAPK1 | MKLN1 | PSMD5 | RBMX2 |
| PNPO | ARID3B | PRPS1 | GJD3 | TECPR1 | CAPS | ZIM2 | H3F3A | POPDC2 | BHLHE22 |

**Table 4:**
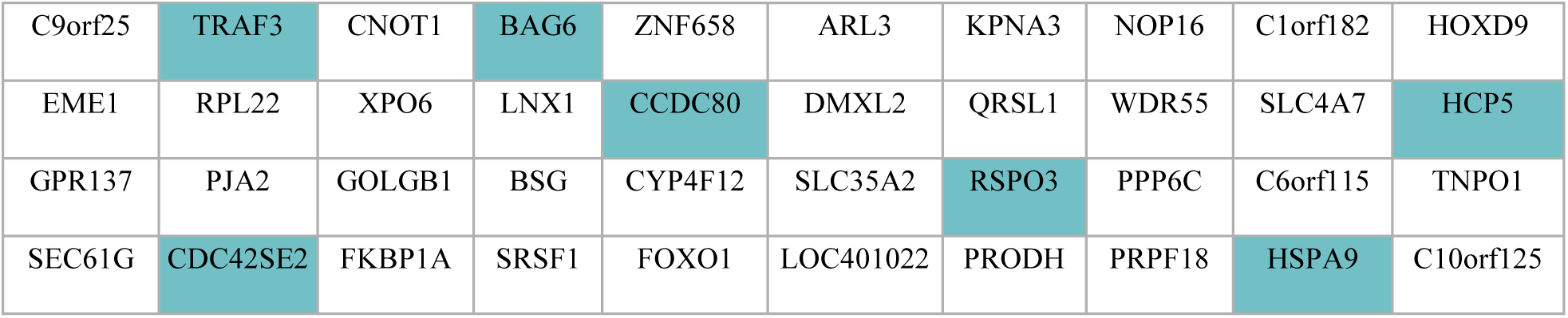
Final unique genes of UC using DGESA: Here the genes highlighted with blue have confirmed their existence in state-of-the-art literature as related with UC. For example, TRAF3 [49], BAG6 [50], CCDC80 [51], CDC42SE2 [52] and HSPA9 [53] are confirmed connection with Inflammatory Bowel Syndrome (IBD) like UC.

## Discussion

The identification of genes implicated in complex diseases is pivotal for advancing our understanding of their underlying biological mechanisms. In this study, we introduce the Differential Gene Expression Based Simulated Annealing (DGESA) algorithm, which employs a novel objective function to enhance the differentiability of gene expression profiles between diseased and normal states. This method addresses a significant gap in existing gene selection strategies by emphasizing the importance of collective differentiability, a concept that has been overlooked in previous research.

Our results from applying DGESA to datasets of Eosinophilic Esophagitis (EoE), Ulcerative Colitis (UC), and Crohn’s Disease (CD) demonstrate its potential in uncovering biologically relevant genes. By comparing the outcomes of DGESA with the Ranked Variance (RV) approach, we found a noteworthy overlap, particularly in the EoE case study, where 10 common genes were identified. This convergence underscores the complementary nature of these methods and suggests that the integration of multiple approaches can enhance the robustness of gene selection processes.

The validation of our findings against existing literature further supports the efficacy of DGESA. Specifically, 10 of the 40 genes identified for EoE, 8 for CD, and 7 for UC were corroborated by previous studies, highlighting their biological relevance. For instance, genes such as KRT79, CRISP2, and IL36G in EoE, and CDKN1A and CYBA in CD, have established roles in the pathophysiology of these diseases, which reinforces the credibility of our method. These validated genes also provide valuable targets for future research and potential therapeutic interventions.

The introduction of the Ranked Variance (RV) approach as an initial step in the DGESA process is particularly noteworthy. By prioritizing genes based on their variance, RV offers a biologically plausible preliminary filter that simplifies the subsequent optimization process. This step not only enhances the efficiency of DGESA but also ensures that the selected genes are inherently variable and thus more likely to be differentially expressed between diseased and normal states.

Our framework’s ability to identify significant molecular signatures associated with gastrointestinal diseases holds promise for broader applications. The DGESA method can be adapted to various other diseases and datasets, potentially uncovering critical genes that have been missed by traditional methods. Moreover, the integration of DGESA with other bioinformatics tools and databases could further enhance its utility and accuracy.

Despite the promising results, several limitations should be acknowledged. The reliance on existing literature for validation, while necessary, may introduce bias, as genes not yet studied or published may be equally important but remain unrecognized. Additionally, the performance of DGESA should be evaluated on larger and more diverse datasets to ensure its generalizability across different populations and disease contexts.

Future research should focus on refining the objective function and exploring alternative strategies for initial gene prioritization. Integrating additional layers of biological data, such as protein-protein interactions and pathway analyses, could provide a more comprehensive understanding of the selected genes’ roles in disease. Furthermore, experimental validation of the identified genes through laboratory studies will be crucial in confirming their functional relevance and potential as therapeutic targets.

## Conclusion

The identification of gene sets that collectively cause a disease, known as the gene selection problem, is a critical area of study in understanding complex diseases. This research introduces two innovative approaches for gene selection in the context of various gastrointestinal diseases: the RV method and DGESA. The RV method prioritizes genes based on their variance, providing an initial perspective on potential biomarkers by identifying genes with significant variability in expression between diseased and normal samples. DGESA, on the other hand, utilizes the principles of simulated annealing to integrate differential gene expression data, refining the selection process by iteratively optimizing gene sets to maximize their discriminative power.

Through a focused case study on EoE and other gastrointestinal diseases like CD and UC, we systematically compare the outcomes of both methods. The RV method initially identifies genes with high variance, offering a broad overview of potential candidates. In contrast, DGESA fine-tunes this selection by incorporating a fitness function that assesses the difference in mean gene expression between diseased and normal states, thus honing in on genes with the most significant impact.

Our results reveal a notable intersection between the two methods, with 10 common genes identified in EoE, highlighting their complementary nature and the robustness of the selection process. Further validation analyses demonstrate that 10 out of the 40 final genes identified by DGESA for EoE are confirmed by existing literature, showcase their biological relevance and potential role in disease pathogenesis. Similarly, in the contexts of UC and Crohn’s Disease CD, 8 and 7 genes, respectively, from the final 40 identified by DGESA are supported by literature evidence, indicating their significance in these diseases. KRT79, CRISP2, IL36G, SPRR2B, SPRR2D, and SPRR2E are among the ten confirmed genes for EoE. NPDC1, SLC2A4RG, LGALS8, CDKN1A, XAF1, and CYBA are included in the literature-confirmed CD set. TRAF3, BAG6, CCDC80, CDC42SE2, and HSPA9 are among the validated genes in the UC final gene collection.

These findings underscore the efficacy of both RV and DGESA in elucidating molecular signatures associated with gastrointestinal diseases. The complementary strengths of the RV method and DGESA provide a robust framework for identifying key genetic contributors to disease, enhancing our understanding of disease mechanisms, and identifying potential therapeutic targets. By integrating these approaches, we can more accurately pinpoint the genes that play pivotal roles in disease development, paving the way for advancements in diagnostics and personalized medicine.

## Supporting information

All Figures

## Data Availability

All data produced in the present study are available upon reasonable request to the authors

