## Supplementary material for "A New Differential Gene Expression Based Simulated Annealing for Solving Gene Selection Problem: A Case Study on Eosinophilic Esophagitis and Few Other Gastro-Intestinal Diseases": All Figures

### Supplement

When the RV approach was applied to the CD vs. normal dataset, informative findings on the variation in gene expression between samples were obtained. It was found that about 40 genes showed decreasing variance by plotting the curve (see Supplement 1), where the x-axis represents the gene index and the y-axis reflects the associated variance. When comparing CD samples to normal samples, this observation points to a significant decrease in the diversity of these genes' expression levels. In the context of CD pathophysiology, this trend of declining variance might point to a level of regulatory homogeneity or a persistent downregulation of gene expression within this subset of genes. The aforementioned results underscore the possible importance of these genes in supporting the molecular processes that underlie the onset and advancement of CD.

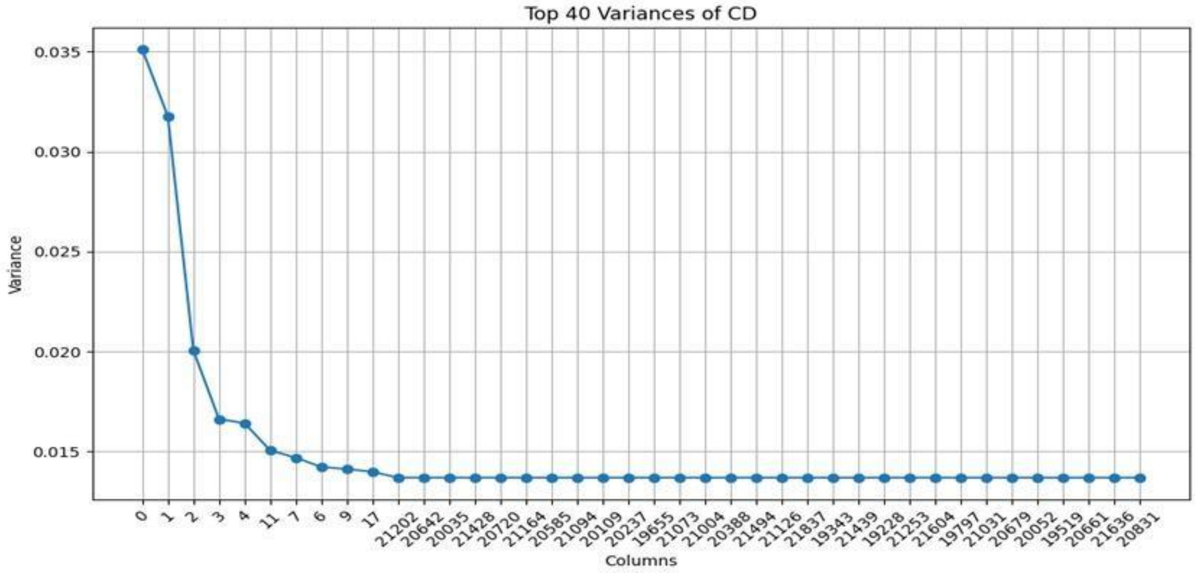


*Supplement 1: Variance of Top 40 Genes in CD vs. normal Gene Expression Data*: The variance of the top 40 genes in the CD gene expression dataset is displayed as a line plot in this figure. The genes with the highest variation are represented by their indices on the x-axis, and their variance values are displayed on the y-axis. The graphic shows that the variance of the first few genes is very high and then flattens off for the next genes.

Using the RV approach on the UC versus normal dataset provided important new information on the variation in gene expression between samples. About 40 genes were found to have a decrease in variance when the curve (see Figure 4) was plotted with the gene index on the x-axis and the matching variance on the y-axis. This suggests that when UC samples are compared to normal samples, there is a notable decrease in the variability of expression levels for these genes. In the context of UC disease, such a decrease in variance points to regulatory homogeneity or continuous downregulation of gene expression within this gene subset. These results highlight the possible significance of these genes in the molecular pathways underlying the onset and course of UC.

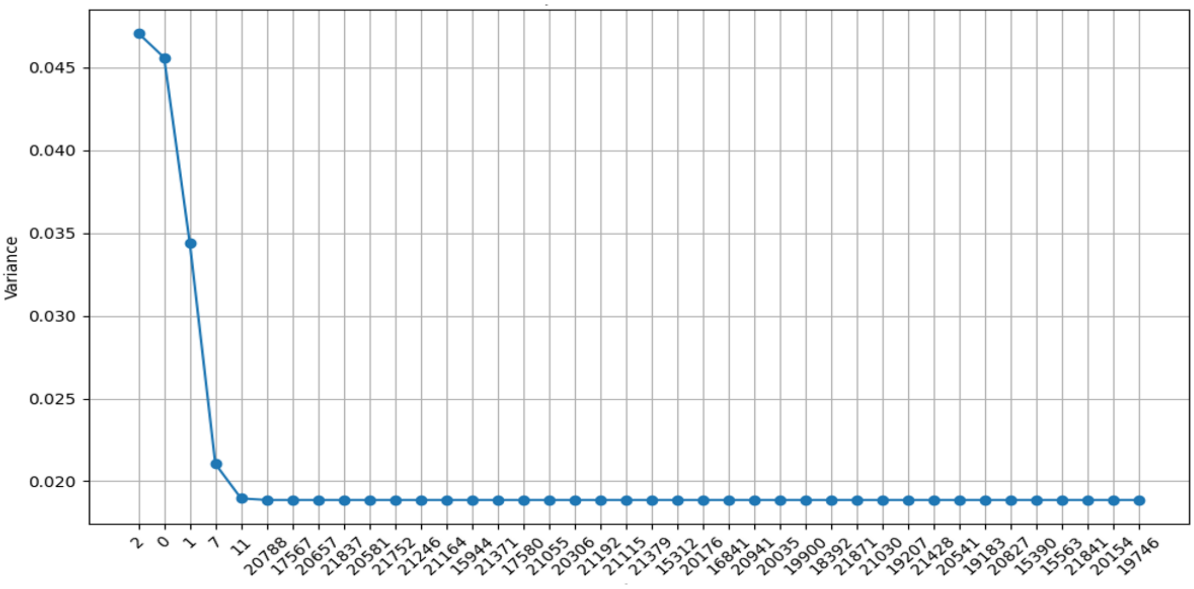


*Supplement 2: Variance of Top 40 Genes in UC Gene Expression Data: The gene expression dataset's variance for the top 40 genes is plotted as a line graph in this picture. The y-axis shows the variance values, while the x-axis shows the indices of the genes with the highest volatility. Plotting shows that a very high variation is seen in the first few genes and then flattens out for the next genes.*

Iterations or steps completed during the simulated annealing optimization process is shown by the x-axis of this curve. Concurrently, the y-axis represents the equivalent fitness values, which measure the potential solutions' efficacy at every iteration. The observed convergence of the curve shows that the fitness values progressively stabilize or improve as the optimization moves through iterations, ultimately arriving at an optimal or nearly optimal solution. The efficacy of DGESA in iteratively optimizing gene selection to maximize its discriminative power between CD and normal samples is indicated by this convergence phenomena. The convergence curve illustrates DGESA's potential as a useful tool for biomarker development and shows how reliable and effective it is at finding disease-associated genetic signatures (see Supplement 3).

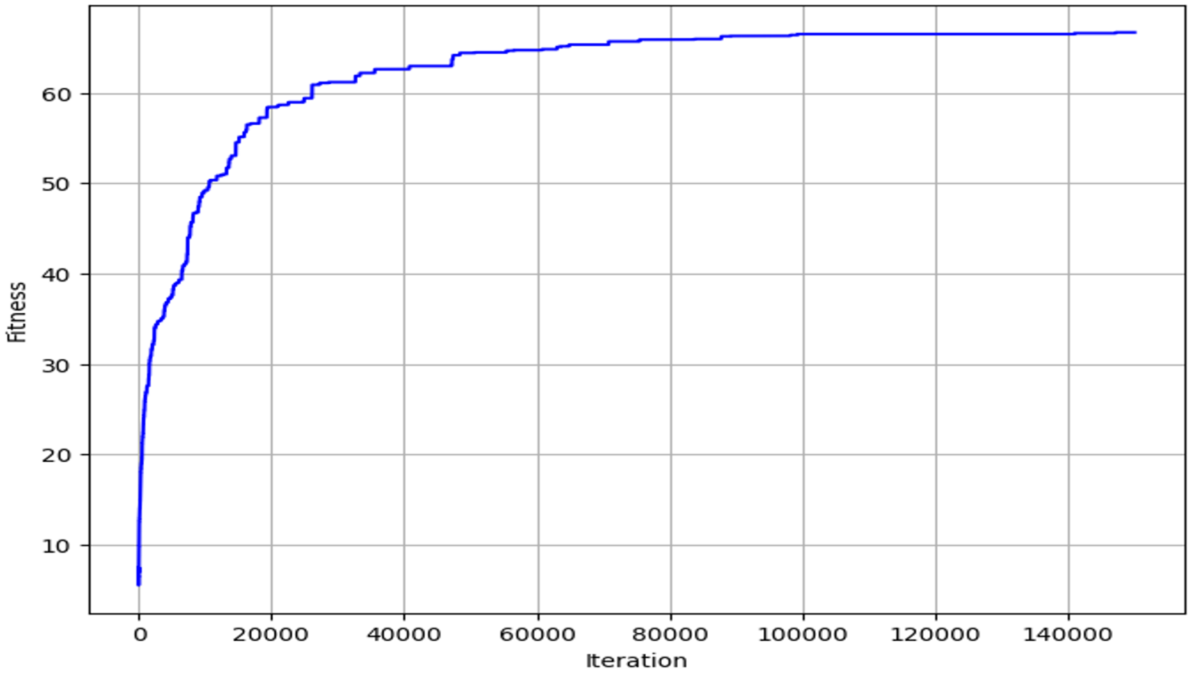


*Supplement 3: Optimization curve of DGESA applied on CD vs. Normal dataset:* Method of Optimization for Finding 40 Discriminative Genes in CD: The optimization procedure used to identify a collection of 40 genes that together enhance the gene expression dataset's discriminating between CD and control samples is depicted in this picture. The optimization process is depicted by the curve, which stabilizes at about 90,000 epochs.

The x-axis of this graph represents the number of iterations or steps finished in the simulated annealing optimization process. Simultaneously, the y-axis displays the corresponding fitness values, which indicate the effectiveness of the possible solutions at each iteration. The curve's observed convergence demonstrates how, as the optimization proceeds through iterations and eventually finds an optimal or nearly optimal solution, the fitness values gradually stabilize or improve. This convergence phenomenon suggests that DGESA is effective in iteratively refining gene selection to maximize its discriminative power between UC and normal samples. The convergence curve (refer to Supplement 4) demonstrates the potential utility of DGESA as a tool for biomarker discovery, as well as the accuracy and consistency of its ability to identify genetic signatures linked with disease.

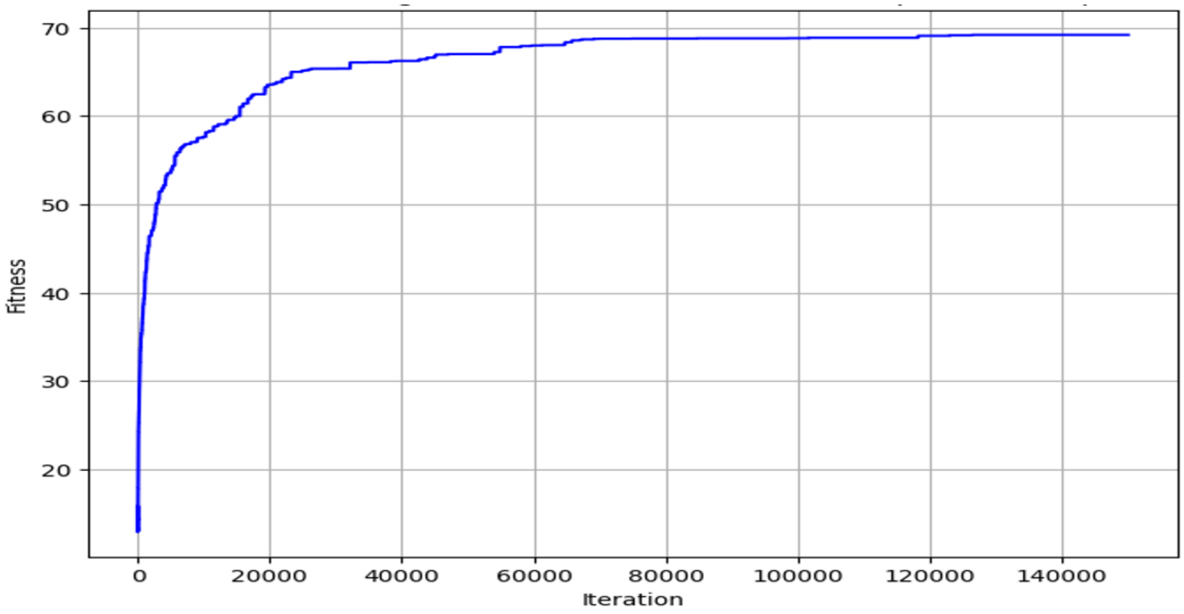


*Supplement 4: Optimization curve of DGESA applied on UC vs. Normal dataset:* 40 Discriminative Genes in UC were found using an optimization process. The optimization procedure to identify a subset of 40 genes that together optimize the discrimination between the UC and control samples in the gene expression dataset is depicted in this image. The graph illustrates how the optimization process is convergent and stabilizes at about 70,000 epochs.
